# Statistics associated with the lethality of COVID-19 by age group and gender in Mexico

**DOI:** 10.1101/2020.06.28.20142117

**Authors:** Carlos Hernandez-Suarez, Efren Murillo-Zamora

**Affiliations:** Facultad de Ciencias, Universidad de Colima, Bernal Diaz del Castillo 340, Colima, Colima, 28040, MEXICO; Departamento de Epidemiología, Unidad de Medicina Familiar No. 19, IMSS, Av. Javier Mina 301, 28000, Colima, Colima. MEXICO

**Keywords:** COVID-19, SARS-CoV-2, IFR, Lethality, Age, Gender, Adjusted lethality

## Abstract

We analyzed outcomes of 102,985 SARS-CoV-2 confirmed cases of patients attending the IMSS (Mexican Institute for National Insurance) from January 2 to August 3, 2020. We calculated relative IFR by age group and gender and introduced the concept of *adjusted lethalities*, that can be used to project the burden of the disease for a population with different demographic characteristics.

## 1. Introduction

Since it was first identified in pneumonia patients in Wuhan, China, the coronavirus disease 2019 (COVID-19) by severe acute respiratory syndrome coronavirus 2 (SARS-CoV-2) has been characterized by its fast-spreading and high related disease burden [1]. The spectrum of symptoms is wide and ranges from asymptomatic infection to severe respiratory illness [2]. By August 4, 2020, almost 20 million confirmed cases of COVID-19 had been reported globally and a fatal outcome was registered in nearly a half million of them [3]. Gender and age-related differences in disease severity and risk of death have been documented [4].

The lethality of a disease, also known as Infection Fatality Rate (IFR) is the fraction of infected individuals that die from the disease. It’s estimation is of the upmost importance for the design of and application of containment measures and to decide if lockdowns are implemented as when these should be reinforced or lifted. Unfortunately, our data does not come from a random sample and we cannot estimate the IFR, instead, we estimate the relative share of the IFR by age group and gender and introduce the concept of *adjusted lethalities* that can be used to project the burden of the disease for a population whose demographic characteristics is known.

### Methodology

We used the database of the Instituto Mexicano de Seguro Social (IMSS, acronym for Mexican Institute for National Insurance) with confirmed SARS-CoV-2 cases. The IMSS provides health services to about 10% of the population in Mexico, over 12 million persons. The database on SARS-CoV-2 we used contains confirmed cases from January 2 to June 23, 2020. We included only cases with disease outcome as dead or recovered, leaving a database with 102,985 cases, from which 55 % were males and 45% females.

When a patient is diagnosed with COVID-19, it follows one of the paths depicted in Figure 1. With this data we constructed Table 1 after dividing the population in seven age categories and two genders.

**Figure 1:**
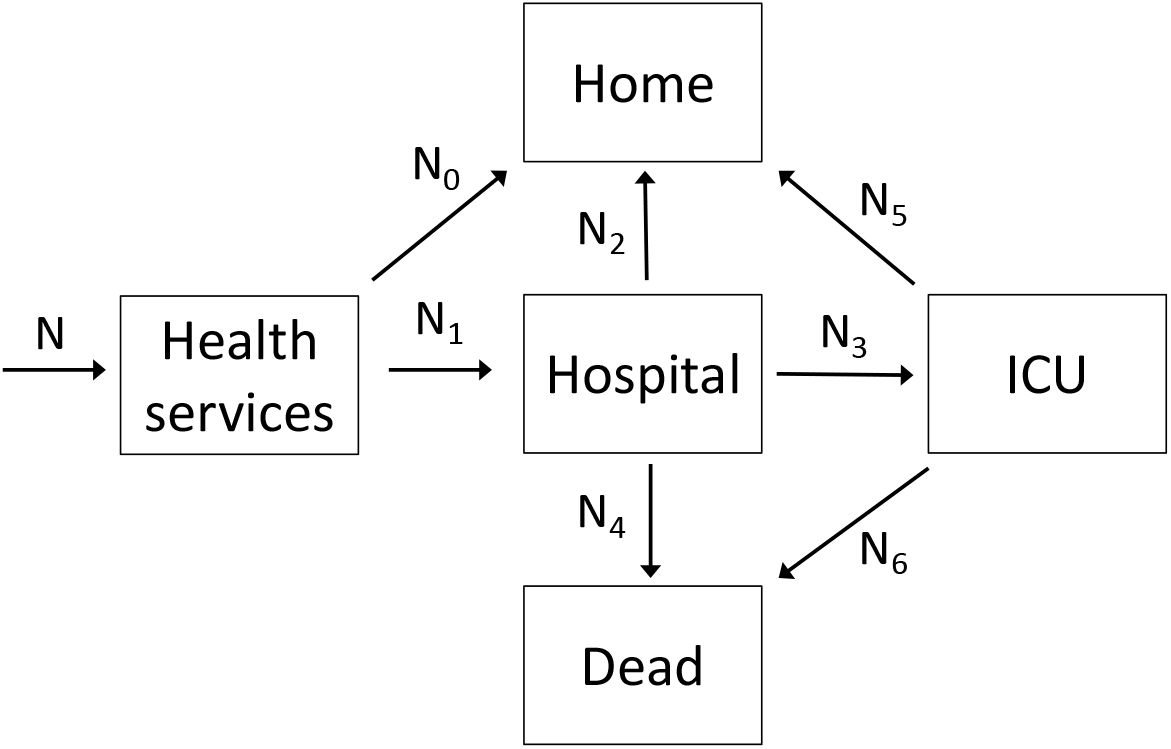
Transitions in our model: *N*_0_*+ N*_1_*= N, N*_2_ + *N*_3_+ *N*_4_ *= N*_1_ + *N*_5_*= N*_3_

**Table 1:** Number of transitions between compartments in Figure 1 by age group and gender.

| **Males** |  |  |  |  |  |  |  |  |
| --- | --- | --- | --- | --- | --- | --- | --- | --- |
| **Age** | **$N$** | **$N_0$** | **$N_1$** | **$N_2$** | **$N_3$** | **$N_4$** | **$N_5$** | **$N_6$** |
| 0-9 | 256 | 97 | 159 | 116 | 20 | 23 | 14 | 6 |
| 10-19 | 367 | 261 | 106 | 83 | 6 | 17 | 3 | 3 |
| 20-29 | 5,627 | 4,585 | 1,042 | 852 | 8 | 182 | 6 | 2 |
| 30-39 | 10,829 | 7,706 | 3,123 | 2,289 | 29 | 805 | 14 | 15 |
| 40-49 | 11,788 | 6,153 | 5,635 | 3,539 | 65 | 2,031 | 32 | 33 |
| 50-59 | 11,566 | 3,799 | 7,767 | 3,914 | 65 | 3,788 | 24 | 41 |
| 60 + | 16,392 | 2,599 | 13,793 | 4,407 | 111 | 9,275 | 27 | 84 |
| Total | 56,825 | 25,200 | 31,625 | 15,200 | 304 | 16,121 | 120 | 184 |

| **Females** |  |  |  |  |  |  |  |  |
| --- | --- | --- | --- | --- | --- | --- | --- | --- |
| **Age** | **$N$** | **$N_0$** | **$N_1$** | **$N_2$** | **$N_3$** | **$N_4$** | **$N_5$** | **$N_6$** |
| 0-9 | 205 | 90 | 115 | 81 | 17 | 17 | 14 | 3 |
| 10-19 | 423 | 316 | 107 | 80 | 3 | 24 | 2 | 1 |
| 20-29 | 5,778 | 4,972 | 806 | 684 | 15 | 107 | 13 | 2 |
| 30-39 | 9,894 | 8,211 | 1,683 | 1,330 | 29 | 324 | 23 | 6 |
| 40-49 | 10,128 | 6,868 | 3,260 | 2,260 | 22 | 978 | 8 | 14 |
| 50-59 | 8,274 | 3,515 | 4,759 | 2,755 | 33 | 1,971 | 13 | 20 |
| 60 + | 11,458 | 2,068 | 9,390 | 3,600 | 54 | 5,736 | 12 | 42 |
| Total | 46,160 | 26,040 | 20,120 | 10,790 | 173 | 9,157 | 85 | 88 |

Table 2 shows a processing of the information in Table 1. Vectors **h**, **c**, **d** and **d**_c_ in Table 2 are described as follows:

**h** = relative frequency of hospitalized patients. **c** = fraction requiring ICU among those that went into hospitalization. **d** = fraction dying among those that went into hospitalization. **d**_c_ = fraction dying among those that went into ICU.

**Table 2:** Statistics associated with the transition of COVID-19 patients between compartments in Figure 1. **h** = distribution of patients arriving to hospitals. **c** = probability of requiring ICU among those that went into hospitalization; **d** = probability of dying among those that went into hospitalization; **d**c = probability of dying among those that went into ICU.

| **Males** |  |  |  |  |
| --- | --- | --- | --- | --- |
| **Age** | **h** | **c** | **d** | **d<sub>c</sub>** |
| 0-9 | 0.0031 | 0.1258 | 0.1824 | 0.3000 |
| 10-19 | 0.0020 | 0.0566 | 0.1887 | 0.5000 |
| 20-29 | 0.0201 | 0.0077 | 0.1766 | 0.2500 |
| 30-39 | 0.0604 | 0.0093 | 0.2626 | 0.5172 |
| 40-49 | 0.1089 | 0.0115 | 0.3663 | 0.5077 |
| 50-59 | 0.1501 | 0.0084 | 0.4930 | 0.6308 |
| 60 + | 0.2666 | 0.0080 | 0.6785 | 0.7568 |

| **Females** |  |  |  |  |
| --- | --- | --- | --- | --- |
| **Age** | **h** | **c** | **d** | **d<sub>c</sub>** |
| 0-9 | 0.0022 | 0.1478 | 0.1739 | 0.1765 |
| 10-19 | 0.0021 | 0.0280 | 0.2336 | 0.3333 |
| 20-29 | 0.0156 | 0.0186 | 0.1352 | 0.1333 |
| 30-39 | 0.0325 | 0.0172 | 0.1961 | 0.2069 |
| 40-49 | 0.0630 | 0.0067 | 0.3043 | 0.6364 |
| 50-59 | 0.0920 | 0.0069 | 0.4184 | 0.6061 |
| 60 + | 0.1815 | 0.0058 | 0.6153 | 0.7778 |

As mentioned before, it is not possible to estimate the IFR from the data available (Table 1) since the death rates are conditional on patients mostly symptomatic and thus they are not a representative sample of the response to an infection. Nevertheless, there is a couple of things that we can obtain from Table 2. The fist one is the *relative lethality* and the second *adjusted lethality*.

## 2. Relative lethality

Table 2 tells us, for instance, that the probability that a female person in age group 40-49 that is hospitalized from infection with SARS-CoV-2 dies is 0.3548, but this does not provides information on the probability that a female infected person in age group 40-49 dies from the disease. Nevertheless, observe that if *f_ij_* is the fraction in the population of individuals of gender *i, i =* 1,2 and group age *j,j =* 1, 2,…, 7., and we assume that everyone is equally likely to become infected but there is a differential response to the disease, then, if *p* is the fraction of individuals in the population that is infected and *β_ij_* is the fraction of infected individuals of gender *i* and age group *j* that become hospitalized, then:

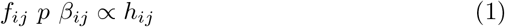

Where *h_ij_* is the fraction of hospitalized that belong to age group *i* and gender *j*. Now, if *α_ij_* is the fraction of individuals hospitalized that die from COVID-19, then

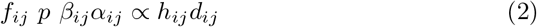

where *d_ij_* is the fraction of individuals of these hospitalized that dies.

Let *θ_ij_* be the *lethality* or infection fatality rate of gender *i* and age group *j*, then, the lethality of gender *i* and age group *j* can be written as:

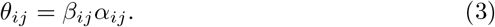

Thus, the IFR by gender and group age follows this relationship:

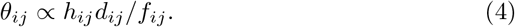

The reason why we only establish that *θ_ij_* is proportional to some value is because we ignore the value of *p*, the true proportion of infected individuals in the population. Thus, we define 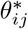 as the *relative lethalitiy*:

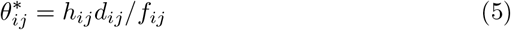

Even although the value of *p* is not known, the ratio of any two relative lethalities 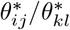 reflects the ratio of the true lethalities, that is:

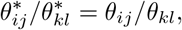

therefore this quotient can be used to compare the lethality of any two groups. Table 3 shows in the last column the relative infection fatality rates in (5), after normalization. In this table f*_M_* and f*_F_* are the relative frequencies by age group and gender in the Mexican population. In this table we can see, for instance, that the lethality of COVID-19 in age group 60+ is 0.37511/0.19795 = 1.9 times higher in males than in females, and that the lethality in the 60+ group compared to the group with less than 20 years age is:

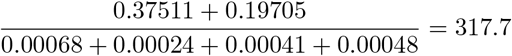

times larger. Table 3 also shows that the lethality among people 50 years or older is about 6 times larger than among people less than 50 years old.

**Table 3:** Normalized relative lethalities. “o” and “Ø” are the Hadamard element-by-element multiplication and division respectively.

| Males |  |  |  |  |
| --- | --- | --- | --- | --- |
| Age | $\mathbf{f}_M$ | $\mathbf{h}$ | $\mathbf{d}$ | $\theta_M^*$ |
| | | | | $\mathbf{h} \circ \mathbf{d} \oslash \mathbf{f}_M$ |
| 0-9 | 0.09871 | 0.00307 | 0.18239 | 0.00052 |
| 10-19 | 0.09976 | 0.00205 | 0.18868 | 0.00036 |
| 20-29 | 0.08130 | 0.02014 | 0.17658 | 0.00403 |
| 30-39 | 0.07203 | 0.06035 | 0.26257 | 0.02029 |
| 40-49 | 0.05566 | 0.10890 | 0.36628 | 0.06609 |
| 50-59 | 0.03851 | 0.15010 | 0.49298 | 0.17722 |
| 60 + | 0.04218 | 0.26656 | 0.67853 | 0.39548 |

| Females |  |  |  |  |
| --- | --- | --- | --- | --- |
| Age | $\mathbf{f}_F$ | $\mathbf{h}$ | $\mathbf{d}$ | $\theta_F^*$ |
| | | | | $\mathbf{h} \circ \mathbf{d} \oslash \mathbf{f}_F$ |
| 0-9 | 0.09577 | 0.0022 | 0.1739 | 0.00037 |
| 10-19 | 0.09824 | 0.0021 | 0.2336 | 0.00045 |
| 20-29 | 0.08709 | 0.0156 | 0.1352 | 0.00223 |
| 30-39 | 0.07908 | 0.0325 | 0.1961 | 0.00744 |
| 40-49 | 0.06096 | 0.0630 | 0.3043 | 0.02900 |
| 50-59 | 0.04225 | 0.0920 | 0.4184 | 0.08399 |
| 60 + | 0.04846 | 0.1815 | 0.6153 | 0.21252 |

## 3. Adjusted lethality

It is possible to estimate the true values of *θ_M_* and *θ_F_*. The lethality of the disease is the probability that a person at random that has been infected with SARS-CoV-2 dies, which is:

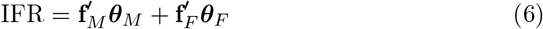

which assumes all individuals are equally likely to become infected. If we have an estimate of the lethality of the disease, *θ*, then we can rescale 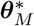 and 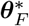 multiplying by a constant *c* so that

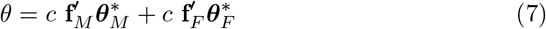

Some estimates of the IFR are in the range 0.05% – 0.25% [5, 6, 7]. Here we will use the lethality reported by [5] of 86 in 100, 000 (about 0.1%) because it is a more recent study with a large sample size, thus, we use *θ* = 0.001. The adjusted lethalities by age group and gender are shown in Table 4.

**Table 4:**
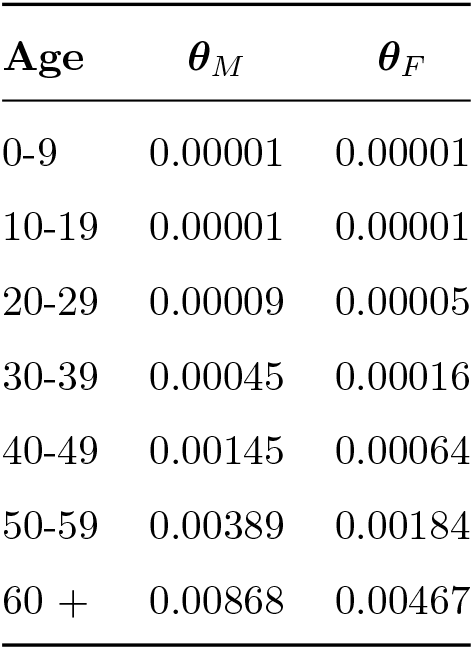
Adjusted lethalities by age group and gender for the Mexican population(*M*=male, *F*=female). The *adjusted lethalities* are obtained from rescaling the *unadjusted lethalities* in Table 3, so that 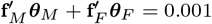.

One immediate application of Table 4 is the possibility of projecting the IFR for another population whose demographics by age and gender are known. Consider for instance the demographics of the Za’atari refugee camp, in Jordan^1^, which is shown in Table 5. We use *θ_M_* and *θ_F_* from Table 4 and the *f_M_* and *f_F_* from the camp’s demographic and thus the IFR projected for this camp is:

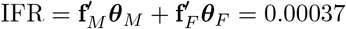

**Table 5:** Population structure of Syrian refugees in Jordan.

| **Age** | $f_M$ | $f_F$ |
| --- | --- | --- |
| 0-9 | 0.1934 | 0.1823 |
| 10-19 | 0.1190 | 0.1116 |
| 20-29 | 0.0714 | 0.0794 |
| 30-39 | 0.0574 | 0.0611 |
| 40-49 | 0.0337 | 0.0348 |
| 50-59 | 0.0127 | 0.0171 |
| 60 + | 0.0110 | 0.0150 |
| Total | 0.4987 | 0.5013 |

Observe this adjusted lethality is about 2.7 times smaller than the overall IFR, which is due to the fact that Za’atari refugee camp is a young population with 3/4 of the refugees being less than 30 years old. Although this comparison used *θ =* 0.001 as the overall IFR of SARS-CoV-2, the observed ratio of 2.7 is independent of this value because the adjusting constant c in 7 cancels out. Clearly, this projection includes only demographic factors and not other health and socioeconomic factors or availability of health services. Figure 2 shows a comparison between the demographics of the Za’atari camp and the *Adjusted lethalities* from Table 4 by age group and gender.

**Figure 2:**
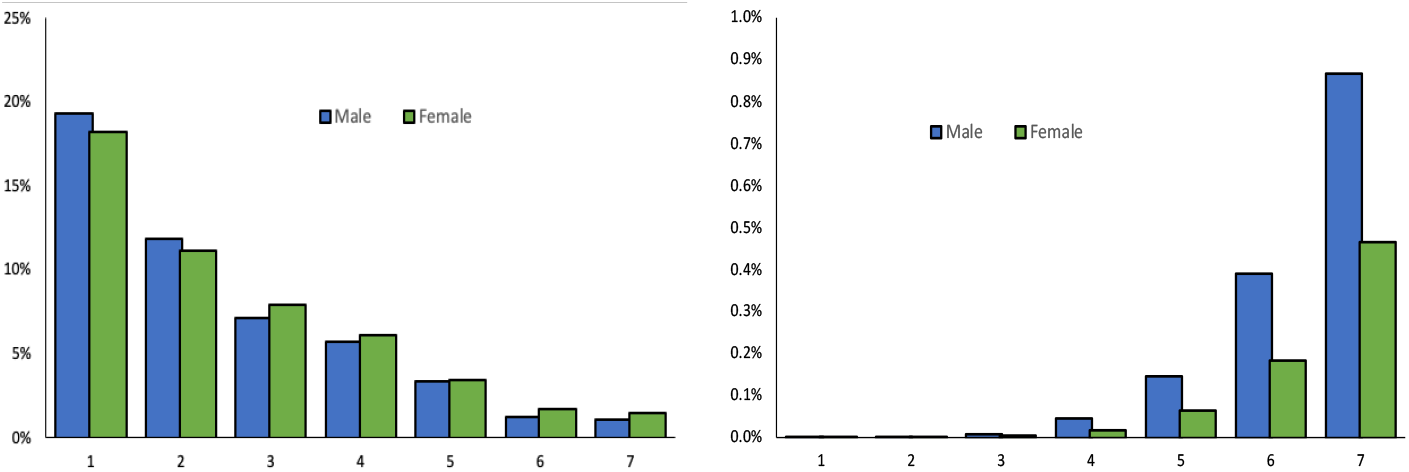
Population frequency in Za’atari camp (left.) and Adjusted lethalities from Table 4 (right) by age and gender. For this particular camp, the lethality affects a group of individuals that are found at a very low frequency in the camp.

### The distribution of deaths across age groups and gender

Another use of the *adjusted lethalities* in Table 4 is the estimation of the distribution of deaths across group ages and genders. The question we want to answer is: what is the probability that a dead individual belongs to a particular gender and age group? This is clearly given by **f***_M_* ○ ***θ****_M_* and **f***_F_* ○ ***θ****_F_* after normalization. For the Mexican population, a plot of this distribution is given in Figure 3.

**Figure 3:**
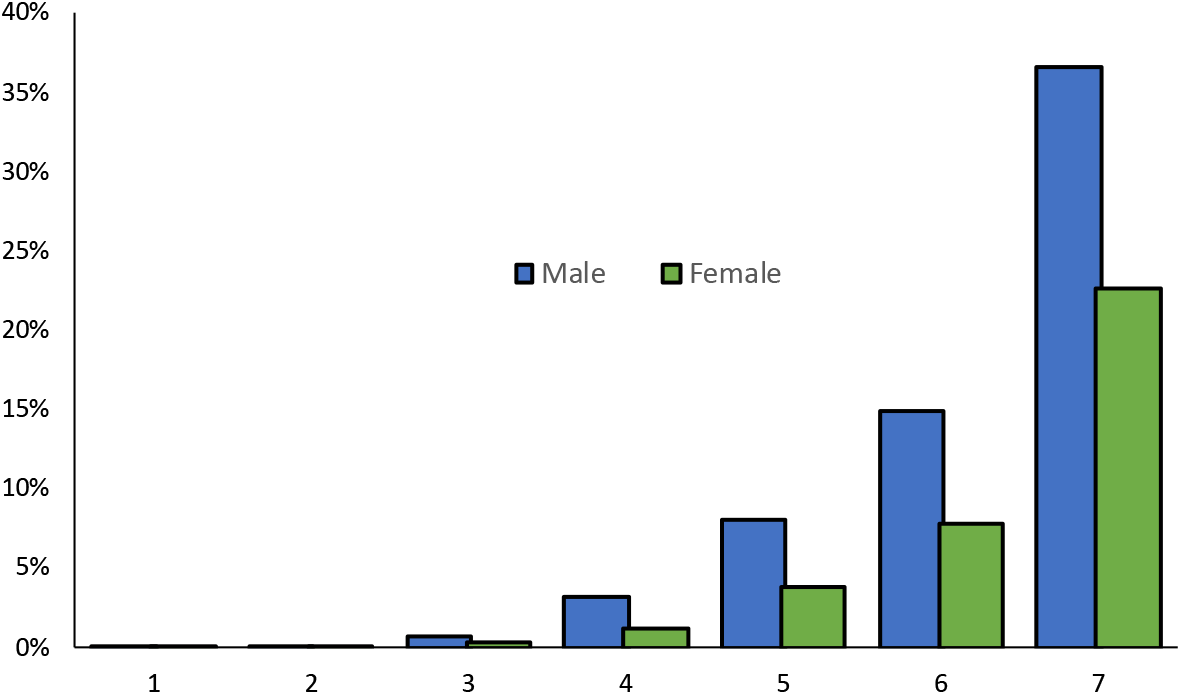
The projected distribution of deaths across age groups and gender for Mexico’s population.

## Data Availability

The data set is only available to researchers of the IMSS for their analysis and exploration.

## Funding and study approval

This work is part of the program “Building the Evidence on Protracted Forced Displacement: A Multi-Stakeholder Partnership”. The program is funded by UK aid from the United Kingdom’s Department for International Development (DFID), it is managed by the World Bank Group (WBG) and was established in partnership with the United Nations High Commissioner for Refugees (UNHCR). The scope of the program is to expand the global knowledge on forced displacement by funding quality research and disseminating results for the use of practitioners and policy makers. This work does not necessarily reflect the views of DFID, the WBG or UNHCR. This study had approval R-2020-601-07 by the Health Research Ethics Committee (601) of the IMSS.

## Footnotes

1 https://data2.unhcr.org/en/situations/syria/location/53

